# Prevalence of Antibiotic-Resistant Bacteria in the Daya River, Bhubaneswar: An Alarming Call for Sewage Disposal

**DOI:** 10.64898/2025.12.29.25343136

**Authors:** Pramod Kumar Jena, Pradeep Kumar Das Mohapatra, Sayed Modinur Rahaman, Soumitra Das, Durga Prasad Barik

## Abstract

The global increase in microbial infections is strongly linked to the widespread dissemination of antibiotic resistance genes in the environment, posing a serious challenge to public health. In the present study, surface water samples were collected from multiple locations along the Daya River near Bhubaneswar, including sites impacted by untreated municipal sewage discharge. The predominant bacterial isolate was identified as *Acinetobacter baumannii*-exhibited resistance to commonly used antibiotics such as ampicillin and ciprofloxacin. These findings highlight the urgent need for improved sewage treatment and waste management practices to prevent contamination of riverine systems and limit the spread of antibiotic-resistant bacteria.

## Introduction

A substantial number of human diseases stem from ingesting contaminated food and water, with water pollution being a primary driver of these infections (Schwarzenbach *et al*., 2010; WHO, 2022). Key pollutants, including hazardous chemicals and waste from healthcare facilities, sewage, agricultural runoff, and industrial discharges, contribute significantly to water contamination. This contamination is closely linked to high chemical oxygen demand levels and the presence of antimicrobial agents in water systems (Wilson., 2014; Sharma *et al*., 2024). In recent years, the declining effectiveness of antimicrobial drugs has become a pressing global concern due to the rising rates of antimicrobial resistance (AMR) (Laxminarayan *et al*., 2013; Oliveira *et al*., 2024). AMR has emerged as a serious worldwide health threat, complicating the prevention and treatment of infections from bacteria, viruses, and fungi. AMR arises over time through genetic mutations in microbes, often triggered by the extensive and prolonged use of antibiotics (Woodford *et al*., ; 2007WHO, 2020a).As antibiotics, which are essential for treating and preventing infections, become less effective, medical professionals are facing growth challenges in controlling infectious diseases worldwide (WHO, 2020a). The rise in AMR is closely linked to poor water quality, inadequate sanitation, and insufficient hygiene practices. Additionally, the mismanagement of medical, hospital, and municipal waste, along with inefficient wastewater treatment systems, has contributed to recent outbreaks of severe diseases. Human behavior, especially poor hygiene and sanitation practices, continues to play a key role in the global spread of AMR (WHO, 2020b).

Widespread resistance to antibiotics such as ampicillin, tetracycline, streptomycin, ciprofloxacin, and erythromycin has been observed among numerous pathogenic bacteria, posing a substantial global challenge for the prevention and treatment of infectious diseases. This resistance is attributed largely to the overuse and misuse of antibiotics (Gao *et al*., 2012; Laxminarayan *et al*., 2013; Sheridan *et al*., 2018; Cameron-Veas *et al*., 2018 ). The presence of antibiotic-resistant genes has been documented in various studies. Resistance is developed by bacteria through multiple mechanisms, including drug efflux, antibiotic inactivation, and ribosomal modification (Leclercq, 2002; Desjardins *et al*., 2004; Poole, 2005). In both Gram-positive and Gram-negative bacteria, acquired resistance often results from chemical modifications to antibiotic molecules, which interfere with protein synthesis at the ribosome and thereby diminish their effectiveness (Wilson, 2014).

The World Health Organization (WHO) has projected that by 2025, over half of the global population will live in regions experiencing water stress (WHO, 2019). Pollution is estimated to contribute to a decline of more than 5% in the GDP of developing countries. This environmental challenge negatively impacts public health and the economy, while also threatening food security, access to clean drinking water, and biodiversity. Therefore, there is an urgent need to develop affordable strategies to address water pollution, particularly that caused by antimicrobial-resistant bacteria. Several studies have reported the presence of antibiotic-resistant bacteria in river systems across India, including the Ganga and Yamuna rivers, which are heavily impacted by sewage discharge and industrial waste (Lamba *et al*., 2020; Prasad *et al*., 2024). While previous studies have explored antibiotic resistance in clinical settings, limited research has focused on the prevalence of multidrug-resistant bacteria in environmental water bodies, particularly in developing regions like India. The specific role of municipal sewage in fostering AMR within natural water ecosystems remains poorly understood. Several recent studies (e.g., Kumar *et al*., 2020; Rathinavelu *et al*., 2024) have highlighted the increasing detection of antibiotic-resistant pathogens in Indian river systems, emphasizing the urgent need for targeted environmental monitoring. Additionally, reports from the WHO and Indian Council of Medical Research (ICMR) stress the importance of identifying resistance hotspots to develop effective intervention strategies (Bhatia, 2024).

This study aims to identify the primary microbial communities in the waters of the Daya River, along with evaluating microbiological contamination, antibiotic resistance, and sensitivity of bacteria in water samples collected from the river in Bhubaneswar, Orissa, India.

## Materials and Methods

### Sample Collection and DNA Extraction

The planned study focuses on the Daya River Basin (Figure 1), as detailed by Jena *et al*. (2023). The basin is located at latitude 19.8833 and longitude 85.5833. Originating at Saradeipur (near Badahati) in Orissa, India, the Daya River flows as a branch of the Kuakhai River. South of Golabai, the river is joined by the Malguni stream, flowing through the Khurda and Puri districts before discharging into Chilika Lake in the northeast. Approximately 8 kilometers (5 miles) south of Bhubaneswar, the historically significant Dhauli Hills lie along the riverbank, surrounded by open fields and featuring major Ashokan edicts carved into rock near the summit road. This area is also believed to be the site of the Kalinga War. Stretching 37 kilometers, the Daya River is impacted by pollutants introduced through the Gangua Canal, carrying industrial and human waste. Over 1.2 million people rely on the Daya River to meet their water needs.

**Figure 1.**
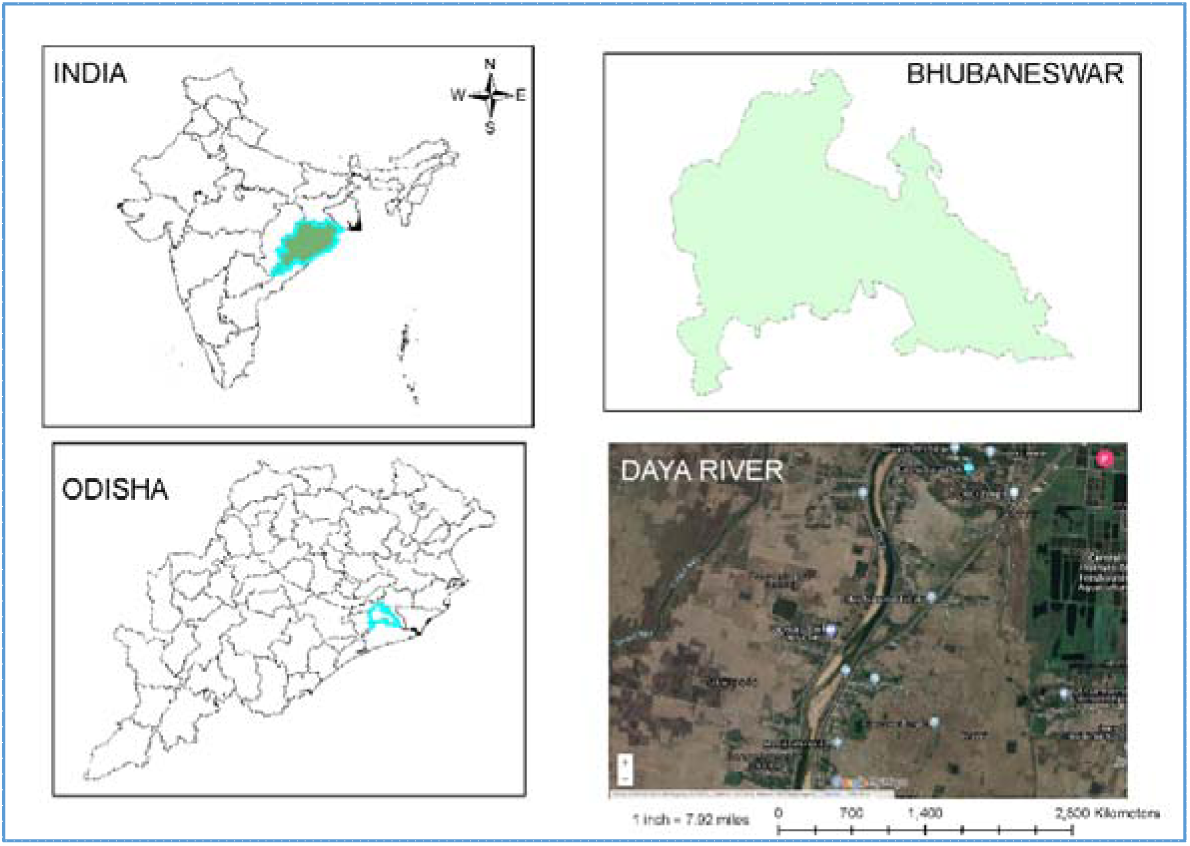
Daya River map taken from Google Earth (Sample collection area)

### Methodology

Water samples were meticulously collected from 8 locations (Table-1) of River Daya, Bhubaneswar, Odisha to ensure a comprehensive representation of the microbial diversity in these environments. These samples were collected using sterile techniques to prevent any contamination, which is crucial for the integrity of downstream molecular analyses (Lane *et al*., 1985). Two hours after the water samples were collected, analysis was done.

**Table-1:** Geo: coordinates of the eight sampling sites of Daya River.

| Sl. No. | Locations | Isolates | Name of the locality | Latitude | Longitude |
| --- | --- | --- | --- | --- | --- |
| 1 | L-1 | D-1 | Daya Bridge, Lingipur | 20°12'32.8"N | 85°51'05.0"E |
| 2 | L-2 | D-2 | Krushnapur | 20°12'37.5"N | 85°50'42.7"E |
| 3 | L-3 | D-3 | Itipur | 20°12'11.1"N | 85°50'06.6"E |
| 4 | L-4 | D-4 | Dola bedi, Palaspursasan | 20°11'23.1"N | 85°49'56.9"E |
| 5 | L-5 | D-5 | Nayahat | 20°13'05.4"N | 85°51'20.2"E |
| 6 | L-6 | D-6 | Jagannathpur | 20°12'41.8"N | 85°50'28.8"E |
| 7 | L-7 | D-7 | Uttarkota | 20°08'47.8"N | 85°49'17.5"E |
| 8 | L-8 | D-8 | Mahirpur | 20°08'18.3"N | 85°49'05.0"E |

**Table 2:** List of Antibiotics and Their Abbreviations used in this present study.

| S.No | Antibiotic | Abbreviation |
| --- | --- | --- |
| 1 | Amoxicillin | AMX |
| 2 | Ceftriaxone | CTR |
| 3 | Colistin | CL |
| 4 | Ciprofloxacin | CIP |
| 5 | Kanamycin | K |
| 6 | Erythromycin | E |
| 7 | Streptomycin | S |
| 8 | Ampicillin | AMP |
| 9 | Cefotaxime | CTX |
| 10 | Ticarcillin | TI |

### Bacterial Isolation

The standard plate count method (Geldreich *et al*., 1972) was employed, wherein 0.1 mL of water samples, which had been serially diluted, was spread on nutrient agar plates (Hi-Media, India). These plates were incubated at 37° C for 24 hours. Following incubation, colonies were examined for their morphological characteristics, and pure cultures were maintained for future study.

### Antimicrobial Susceptibility Testing

Antimicrobial sensitivity testing was conducted using the disk diffusion method on Mueller–Hinton agar (Hi-Media, India). Ten antibiotics, encompassing cephalosporins, β-lactams, sulfonamides, quinolones/fluoroquinolones, macrolides, and aminoglycosides, were evaluated, with details provided in the supplementary materials. Antibiotic concentrations adhered to the Clinical and Laboratory Standards Institute (CLSI, 2019) guidelines. The test organisms were spread onto Mueller–Hinton agar plates (Hi-Media, India) using a lawn culture approach, and antibiotic-impregnated disks were placed on the agar surface. After incubating at 37° C for 24 hours, inhibition zones were measured to classify resistance, intermediate sensitivity, and susceptibility in line with CLSI (2019) standards (Weinstein *et al*., 2020; Reller *et al*., 2009).

### Molecular characterization of multidrug-resistant bacteria

The extraction of high-quality genomic DNA have been completed using the following steps: First added 1 ml of Xploregen gDNA Extraction Buffer™ 1 to a beaded vial containing the sample to break down cell walls and membranes, followed by vortexing for 10 minutes to ensure thorough lysis. Then, 300 µl of Xploregen gDNA Extraction Buffer™ 2 was added and vortexed for 7 minutes to enhance lysis and solubilize proteins. The mixture was centrifuged at 10,000 rpm for 3 minutes, and 800 µl of the supernatant was carefully transferred to a sterile vial. Next, 200 µl of Xploregen gDNA Extraction Buffer™ 3 was added, briefly vortexed, and centrifuged to precipitate proteins. After that 800 µl of the cleared supernatant with 1 ml of Xploregen gDNA Extraction Buffer™ 4 to ensure homogeneity. Subsequently, 700 µl of the lysate was transferred to a spin column and centrifuged at 10,000 rpm to bind the DNA. This step was repeated to ensure maximum DNA binding. Afterward, the column was washed with 600 µl of Xploregen gDNA Extraction Buffer™ 5, followed by Xploregen gDNA Extraction Buffer™ 6 to remove impurities. The column was dried by centrifuging for 5 minutes, and the purified DNA was eluted with 30 µl of Xploregen gDNA Extraction Buffer™ 7. This protocol yielded DNA suitable for molecular analyses like 16S rDNA sequencing, essential for bacterial identification and phylogenetic studies.

### DNA Quantification

The concentration of extracted DNA was determined using spectrophotometric analysis, a reliable and widely used method for nucleic acid quantification (Wilfinger *et al*., 1997). This technique involves measuring the absorbance of DNA at specific wavelengths, typically at 260 nm, where nucleic acids absorb UV light maximally. The purity of DNA was also assessed by calculating the absorbance ratio at 260 nm and 280 nm (A260/A280), which provides an indication of protein contamination (Sambrook & Russell, 2001).

The quantification was performed using a NanoDrop spectrophotometer, which offers precise and quick measurements with minimal sample volume (Desjardins. *et. al.*, 2010). This concentration is sufficient for downstream applications, including PCR amplification and sequencing, ensuring that high-quality genomic material is available for molecular analyses

### PCR Amplification and Sequencing

To ensure accuracy and integrity in the amplification of the 16S rDNA sequences, high-fidelity PCR was employed. High-fidelity polymerases were specifically chosen for their low error rates and high processivity, essential for generating reliable sequence data (Kieleczawa, 2006). The 16S rRNA gene sequences from the predominant antibiotic-resistant isolate (D-1 and D-2) were selected for comprehensive characterization. Amplification of these genes was carried out using polymerase chain reaction (PCR) with universal primers 16SF: 5′-GGATGAGCCCGCGGCCTA-3′ and 16SR: 5′-CGGTGTGTACAAGGCCCGG-3′ under defined conditions. The PCR mixture included 0.01 mol/L Tris-HCl, 0.0015 mol/L MgCl2, 10 ng of genomic DNA, 0.0004 mol/L of each dNTP, 0.5 U of DNA polymerase, 0.05 mmol/L KCl, and 0.1% Triton X-100.

The PCR amplification was carried out under the following optimized cycling conditions: an initial denaturation at 94°C for 3 minutes to fully denature the DNA template and produce single strands for primer binding. This was followed by 30 cycles of denaturation at 94°C for 1 minute to repeatedly separate the DNA strands. Annealing occurred at 50°C for 1 minute, allowing primers to bind to complementary sequences based on their melting temperatures. The extension phase took place at 72°C for 2 minutes, during which DNA polymerase synthesized new strands by adding nucleotides. A final extension step at 72°C for 7 minutes ensured all remaining single-stranded DNA was fully extended to produce complete PCR products. Post-amplification, the PCR products were separated on a 1.8% agarose gel, stained with ethidium bromide, and visualized. The products were then purified to eliminate excess primers, nucleotides, and other reaction components that could interfere with sequencing. Finally, the purified PCR products were sequenced bi-directionally using the ABI 3130xl Genetic Analyzer, a capillary electrophoresis-based platform known for its high accuracy and throughput.

### Bioinformatic Analysis and Phylogenetic Tree Construction

The obtained sequences were subjected to bioinformatics analysis to ensure accurate identification and phylogenetic placement of the isolated strain. The raw sequence data were first cleaned and trimmed to remove any low-quality bases and primer sequences using the software package Trimmomatic (Bolger *et al*., 2014). This step is crucial for improving the quality and reliability of downstream analyses. The obtained 16S rRNA sequences were analyzed through a BLAST search (www.ncbi.nlm.nih.gov/BLAST) using the nBLAST algorithm to identify similar sequences. Phylogenetic analysis was conducted using the neighbor-joining method, and phylograms were constructed (Saitou *et al*., 1987). These phylograms were evaluated through bootstrapping in Clustal X and visualized with TreeView software (Altschul *et al*., 1990). The 16S rRNA gene sequences for isolates D-1 and D-2 were submitted to GenBank (www.ncbi.nlm.nih.gov/genbank).

## RESULTS AND DISCUSSION

### Study Area

Among the eight sampling locations (L-1 to L-8), bacterial isolates D-1 (from L-1) and D-2 (from L-2) were chosen for further molecular characterization. The decision was based on multiple factors observed during primary screening. First, within 24 hours of incubation, these isolates exhibited the most distinct and abundant colony morphologies compared to the others, indicating their dominance in the microbial community at these sites.These isolates displayed higher levels of antibiotic resistance in the preliminary antibiogram, with minimal zones of inhibition (≤0.7 cm) against multiple tested antibiotics, including ampicillin and ciprofloxacin. Because of their prevalence, distinct morphology, and multidrug resistance pattern, D-1 and D-2 were prioritized for detailed genetic identification through 16S rRNA sequencing. This selection strategy ensured that the isolates chosen for molecular analysis represented the most relevant bacterial types in terms of both ecological dominance and public health concern.

### Colony Morphology and Gram Reaction of Bacterial Isolates

A total of eight morphologically different distinct bacterial isolates (D-1 to D-8) were isolated and characterized based on their macroscopic colony morphology and Gram staining. The colonies exhibited a circular shape across all isolates, with notable variations in size (2–4 mm), pigmentation (white to creamy white), texture (smooth to dry), elevation (flat, raised, umbonate), and margin (entire or scalloped) (Table 3).

**Table-3:** Colony morphology and gram reactions bacterial isolates from Daya river.

| Isolate No. | Size (mm) | Shape | Colour | Texture | Elevation | Margin | Gram nature |
| --- | --- | --- | --- | --- | --- | --- | --- |
| D-1 | 3 | Circular | White | Smooth | Raised | Entire | - ve |
| D-2 | 3 | Circular | White | Smooth | Raised | Entire | -ve |
| D-3 | 4 | Circular | White | Smooth | Flat | Scalloped | + ve |
| D-4 | 3 | Circular | Creamy white | Smooth | Umbonate | Entire | -ve |
| D-5 | 2 | Circular | White | Smooth | Raised | Entire | -ve |
| D-6 | 3 | Circular | Creamy white | Dry | Flat | Entire | + ve |
| D-7 | 3 | Circular | White | Smooth | Raised | Entire | -ve |
| D-8 | 2 | Circular | Creamy white | Smooth | Raised | Entire | -ve |

Most isolates, including D-1, D-2, D-5, D-7, and D-8, displayed smooth, raised colonies with entire margins and were Gram-negative, indicative of possible affiliation with non-sporulating Gram-negative genera frequently reported in freshwater systems. Notably, D-3 and D-6 were Gram-positive. DL-3 exhibited a flat colony with a scalloped margin, while D-6 showed a dry, flat colony, with potential spore-forming characteristics, which are typical of the genus Bacillus (Logan & De Vos, 2015). The dry texture of D-6 may also indicate a stress-adaptive phenotype commonly associated with endospore formers in nutrient-limited environments (Nicholson *et al*., 2000).D-4, although similar in color to D-6, differed in its umbonate elevation and smooth surface, remaining Gram-negative. Such variations in elevation and texture may reflect adaptive responses to specific environmental pressures or biofilm-forming potential (O’Toole *et al*., 2000).These morphological and Gram-staining traits serve as fundamental parameters for preliminary bacterial identification. However, molecular techniques such as 16S rRNA gene sequencing are required for accurate taxonomic classification (Janda & Abbott, 2007).

The majority of these isolates were motile bacilli. Their colonies typically displayed a circular shape, varying from small to large, with smooth, well-defined edges, a convex shape, and a shiny surface. These traits, along with other selective and differential characteristics, were observed on the culture media. The primary isolates from each selective and differential medium were labelled D-1 and D-2. The detailed biochemical characteristics of these isolates, determined through various biochemical tests, are summarized in Table 4.

**Table-4:**
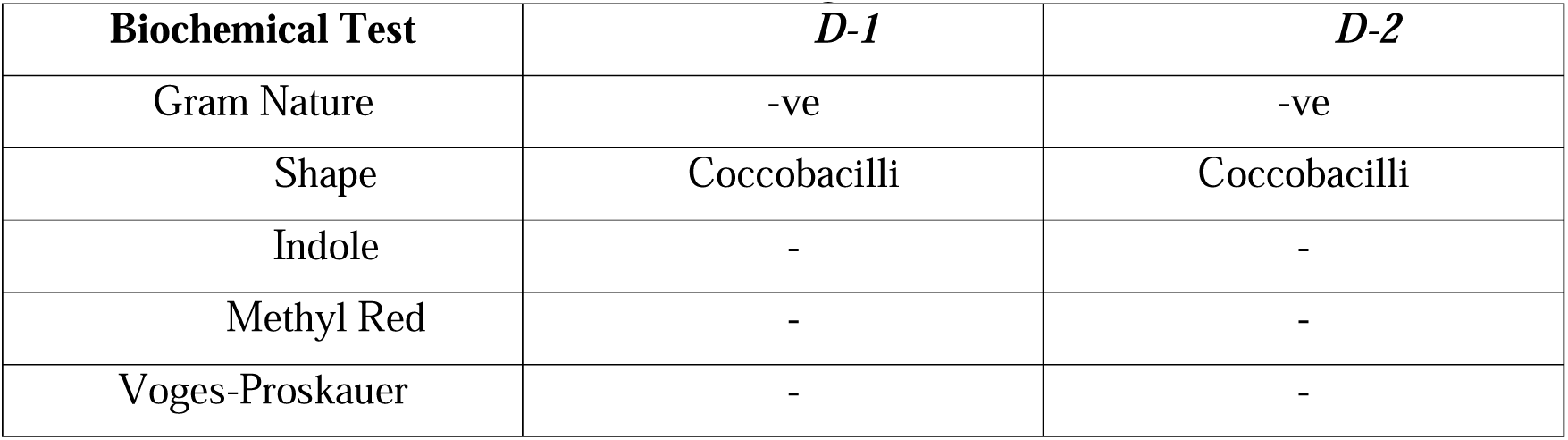
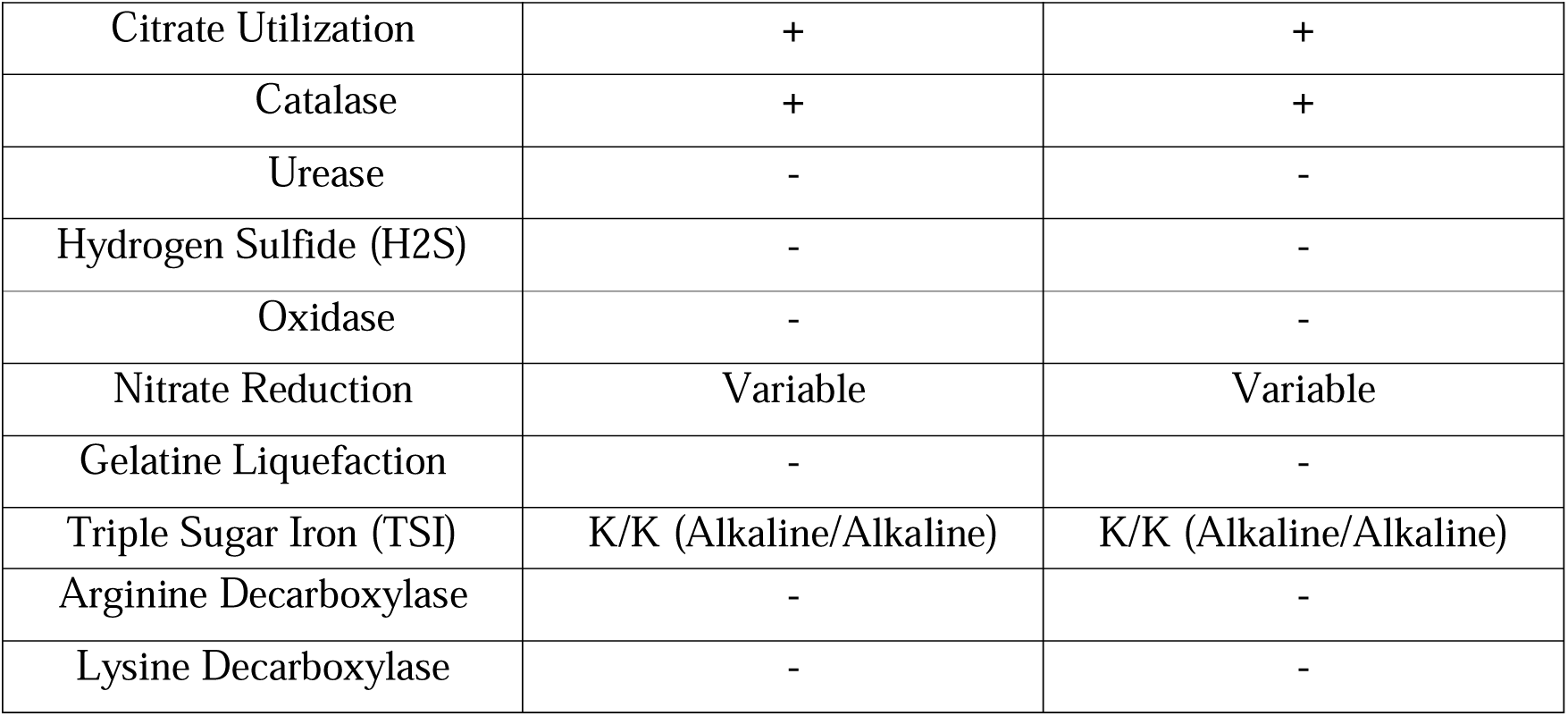
Characteristics of the bacteriological isolates.

### Antimicrobial Susceptibility

The zone of inhibition (ZOI) assay demonstrated varying levels of resistance among the isolates to several antibiotics, including Amoxicillin (AMX), Ceftriaxone (CTR), Colistin (CL), Ciprofloxacin (CIP), Kanamycin (K), Erythromycin (E), Streptomycin (S), Ampicillin (AMP), Cefotaxime (CTX), and Ticarcillin (TI), (Table-4). Additionally, multidrug resistance was observed, underscoring the complexity of antibiotic resistance in aquatic environments, in line with recent research (Das *et al*., 2023). The table presents data on the antimicrobial susceptibility of different isolates from eight locations (Table-1). The measurements reflect the inhibition zones (in mm) for various antibiotics.

The table-5 presents antimicrobial resistance data of the isolates from eight different locations (D-1 to D-8) across 10 different antibiotics: Ampicillin (AMP10), Amoxicillin (AMX), Ciprofloxacin (CIP5), Chloramphenicol (CL10), Ceftriaxone (CTR30), Cefotaxime (CTX), Erythromycin (E15), Kanamycin (K30), Streptomycin (S10), and Tetracycline (TI75). Resistance values range from 0 to 2.7, indicating varying levels of susceptibility. Isolates D-1, D-2, D-4, and D-6 show relatively high resistance to multiple antibiotics, particularly Ciprofloxacin (CIP5) and Kanamycin (K30). In contrast, D-3 and D-5 display lower resistance, with D-5 being more susceptible overall. D-7 shows low or no resistance to some antibiotics, such as Chloramphenicol (CL10), Ceftriaxone (CTR30), and Streptomycin (S10). Meanwhile, D-8 exhibits moderate resistance to several antibiotics but higher resistance to Amoxicillin (AMX) and Kanamycin (K30). This data underscores variability in resistance patterns across different locations and antibiotics, indicating differing antibiotic efficacy in varying environments and micro-organism.

**Table-5:** Antibiogram of isolated bacteria against 10 selected antibiotics.

| Isolate | AMP10 | AMX | CIP5 | CL | CTR30 | CTX | E15 | K30 | S10 | TI75 |
| --- | --- | --- | --- | --- | --- | --- | --- | --- | --- | --- |
| D-1 | 0.3 | 0.4 | 0.5 | 0.2 | 0.3 | 0.4 | 0.1 | 0.3 | 0.2 | 0.4 |
| D-2 | 0.5 | 0.6 | 0.7 | 0.4 | 0.5 | 0.6 | 0.2 | 0.5 | 0.3 | 0.5 |
| D-3 | 1.2 | 1.5 | 2.0 | 1.3 | 1.4 | 1.7 | 1.2 | 1.8 | 1.7 | 1.6 |
| D-4 | 1.5 | 1.7 | 2.2 | 1.6 | 1.8 | 2.0 | 1.5 | 2.0 | 1.8 | 1.7 |
| D-5 | 1.0 | 1.3 | 1.8 | 1.1 | 1.2 | 1.5 | 1.0 | 1.6 | 1.4 | 1.5 |
| D-6 | 1.3 | 1.6 | 2.1 | 1.4 | 1.5 | 1.8 | 1.3 | 1.9 | 1.7 | 1.6 |
| D-7 | 1.1 | 1.4 | 1.9 | 1.2 | 1.3 | 1.6 | 1.1 | 1.7 | 1.5 | 1.5 |
| D-8 | 1.4 | 1.7 | 2.3 | 1.5 | 1.7 | 2.1 | 1.4 | 2.1 | 1.9 | 1.8 |

The antibiogram heatmap (Figure-2) revealed notable spatial variability in the antibiotic susceptibility profiles of bacterial isolates collected from different locations along the Daya River. Isolates D-1 and D-2 exhibited very low zones of inhibition (ranging from 0.1 to 0.7 cm) across all tested antibiotics, indicating a high level of antibiotic resistance among microbial communities at these sites. Such resistance could be attributed to prolonged exposure to antibiotic contaminants or other pollutants typically associated with urban runoff and untreated sewage inputs.

**Figure-2:**
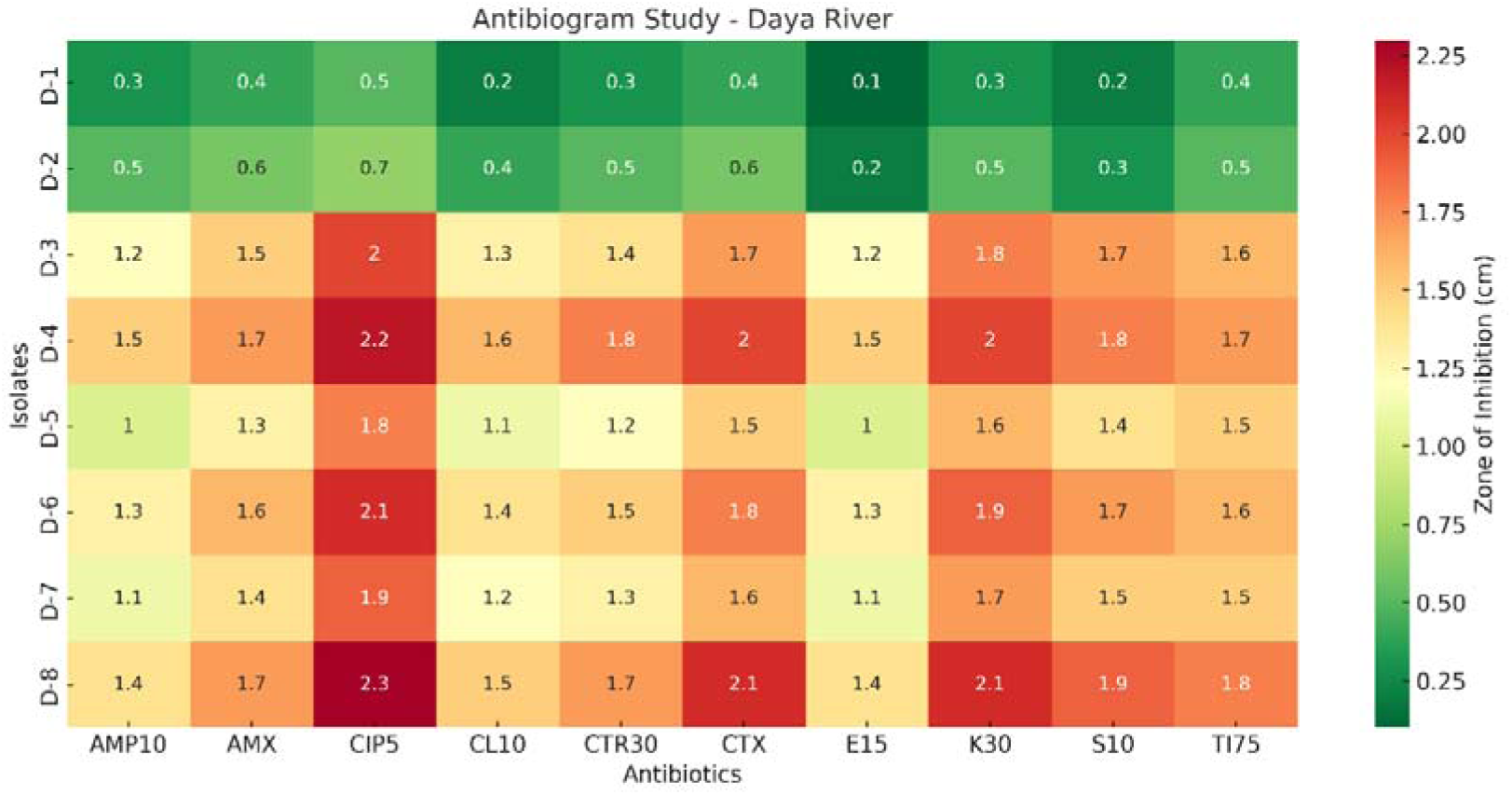
Heatmap of antibiotic susceptibility profiles of bacterial isolates (D-1 to D-8) from eight locations (L1 to L8) along the Daya River. Zones of inhibition (cm) were measured by the disc diffusion method. Green indicates higher resistance, and dark red indicates greater susceptibility.

In contrast, bacterial isolates D-4, D-6, and D-8 showed significantly higher zones of inhibition, particularly against Ciprofloxacin (CIP5) and Kanamycin (K30), with inhibition zones frequently exceeding 2.0 cm. The maximum zone of inhibition (2.3 cm) was recorded on D-8 against Ciprofloxacin, suggesting that bacterial populations at this site were more sensitive to certain antibiotics. The relatively higher susceptibility observed at these locations may reflect lesser or more recent exposure to antibiotic pollutants, or differences in local environmental conditions such as dilution effects, water flow dynamics, or microbial community composition.

Interestingly, intermediate susceptibility patterns were observed on D-3, D-5, and D-7, where inhibition zones ranged between 1.0 and 1.9 cm for most antibiotics. These mixed responses indicate transitional zones where selective pressure from antibiotic contamination might be moderate.

The observed variations across sites underscore the potential impact of anthropogenic activities — such as agricultural runoff containing veterinary antibiotics, discharge from healthcare facilities, and domestic sewage inputs — on shaping the antibiotic resistance landscape within the riverine ecosystem.

### Molecular characterization of bacterial isolates

For this study, the DNA concentration of the isolates from sample location: L1 with isolation code marked as D-1 and sample location L2 with isolation code marked as D-2 were found to be 167 ng/µl and 155 ng/µl respectively. These are selected based on most abundant and dominant types of colonies. The isolates D-1 and D-2, which exhibited most abundant and dominant types of colonies, were characterized through 16S rDNA gene sequencing. The 16S rDNA sequence of the sample (1252bp & 1217bp) underwent rigorous alignment with reference sequences from the NCBI database to accurately determine its phylogenetic relationships. The alignment was performed using the BLAST (Basic Local Alignment Search Tool) algorithm, which is highly effective for identifying homologous sequences by comparing nucleotide sequences against the vast database of known sequences (Altschul *et al*., 1990). This approach ensures precise matching and alignment, which is crucial for accurate phylogenetic analysis. The aligned sequences were then analyzed to construct a phylogenetic tree, which provides a visual representation of the evolutionary relationships among the sequences.The phylogenetic analysis of water samples from isolates D-1 reavels 100% match with *Acinetobacter* species and D-2 reveals the 100% match to *Acinetobacter baumannii*. This species is a prominent pathogen known for its multi-drug resistance and its ability to survive in diverse environmental conditions, making it a critical subject of study.(Janda *et al* 2007)

These results demonstrate the robustness of the phylogenetic analysis and confirm the identity of the isolated strain as *Acinetobacter baumannii*. The use of comprehensive bioinformatics tools and the comparison with a broad range of reference sequences provide a solid foundation for the accurate classification of bacterial strains, which is essential for understanding their genetic diversity and evolutionary history (Thompson *et al*., 1994; Kumar *et al*., 2018).

To elucidate the genetic relationship of the isolated strains from isolates D-1 and D-2, a phylogenetic tree (Fig-3 and 4) was constructed using the Weighbor algorithm. The Weighbor method, which stands for "Weighted Neighbor Joining," is an effective technique for generating phylogenetic trees from molecular data by minimizing the impact of alignment errors and inconsistencies (Bruno *et al*., 2000). This algorithm is particularly useful for handling large datasets and provides robust and reliable tree topologies that reflect the evolutionary relationships among the sequences. The tree reveals that the D-1 and D-2 strain clusters closely with *Acinetobacter baumannii* strain, indicating a high degree of sequence similarity respectively. This suggests that the D-1 and D-2strain is closely related to this reference strain, reflecting a shared evolutionary lineage.

**Figure-3:**
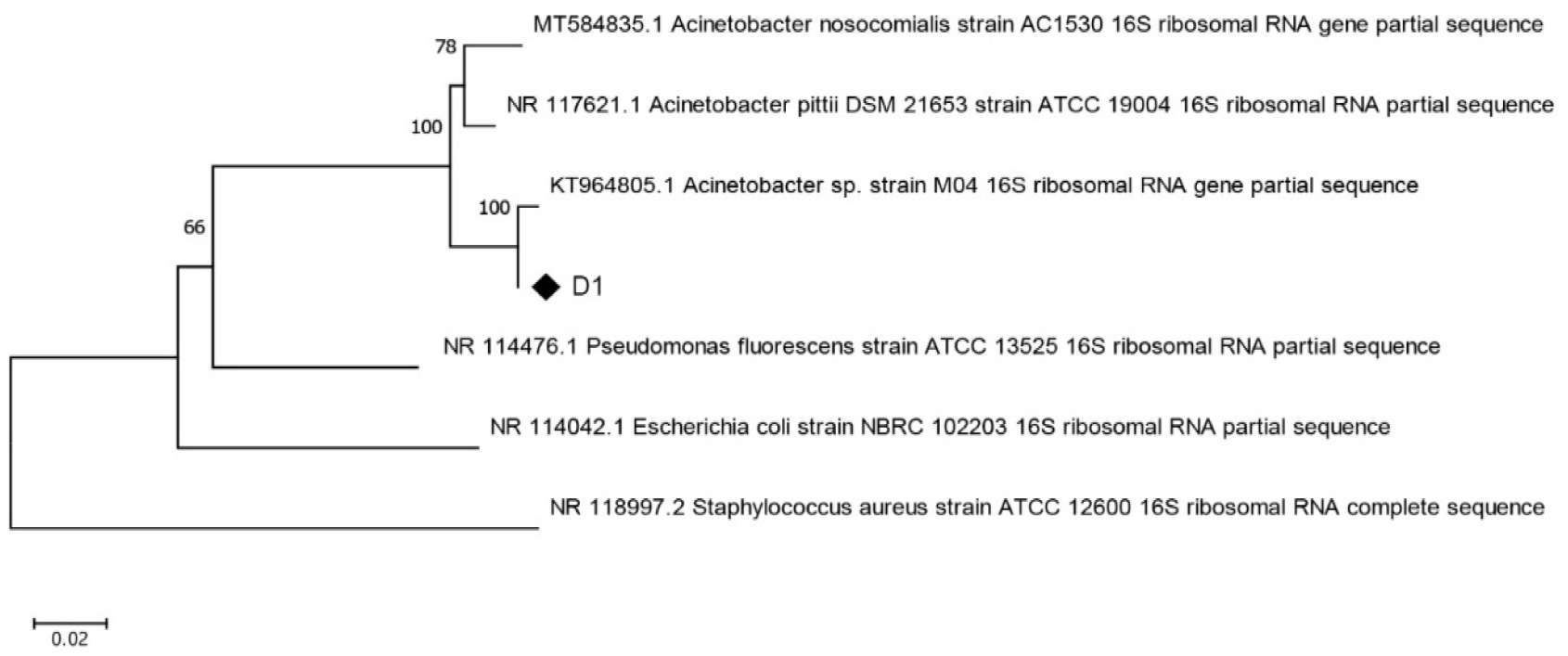
Phylogenetic Tree Showing the Relationship of location D-1 of location L-1with *Acinetobacter* species based on 16S rRNA Gene Sequences

**Figure-4:**
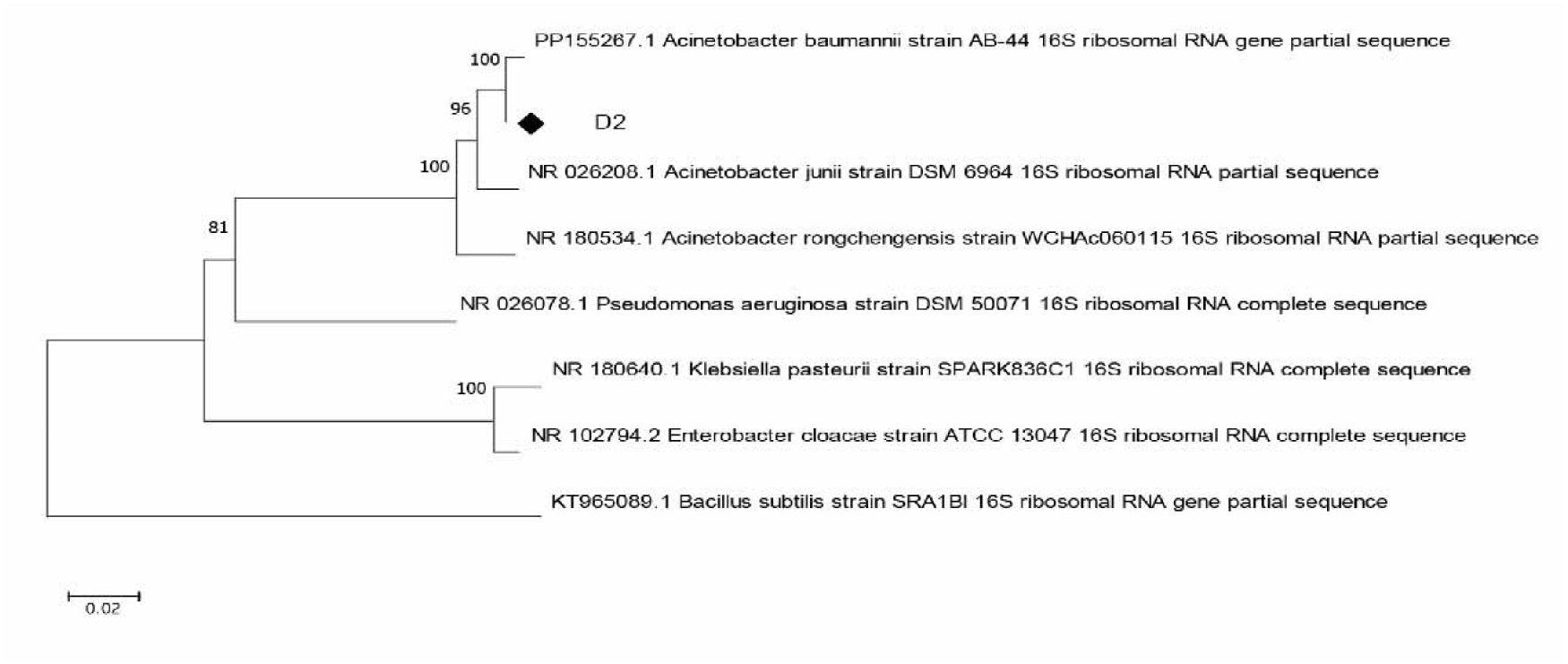
Phylogenetic Tree Showing the Relationship of location D-2 of loation L-2 with *Acinetobacter baumannii* Strains based on 16S rRNA Gene Sequences

### Comparison with Previous Studies on Indian Rivers

The antibiotic resistance patterns observed in the Daya River closely align with trends reported in other major Indian rivers, notably the Ganges and Yamuna. Based on the heatmap analysis, high resistance was detected against Amoxicillin (AMX) and Ampicillin (AMP10), with mean zones of inhibition of 1.32 cm and 0.71 cm, respectively, indicating widespread β-lactam resistance among the isolates. These findings are consistent with earlier studies from the Ganges River, where coliform bacteria exhibited significant resistance to β-lactam antibiotics, including ampicillin, across multiple river stretches (Acharya *et.al.,*2024).

Intermediate susceptibility was observed against Ciprofloxacin (CIP5) in our samples, reflected by a mean zone of inhibition of 1.61 cm. This trend is comparable to findings from the Yamuna River, where ciprofloxacin concentrations in surface water have been reported as high as 1,400 ng/L, correlating with the presence of ciprofloxacin-resistant bacterial populations (Singh *et.al*.,2019).

These comparisons underscore the urgent need for improved sewage treatment infrastructure, as rivers such as the Daya, though smaller in scale than the Ganges or Yamuna, are exhibiting similar resistance trends and could become significant reservoirs for antimicrobial resistance if current practices persist.

## Discussion

The detection and identification of *Acinetobacter baumannii* strains in samples from River Daya, Bhubaneswar, Odisha, highlight a significant concern regarding the widespread distribution of this pathogen beyond traditional hospital settings. *Acinetobacter baumannii* is known for its role as a nosocomial pathogen, exhibiting remarkable resistance to multiple antibiotics and contributing to severe infections in immunocompromised patients (Wong *et al*., 2017). However, its presence in environmental habitats signifies that it is not confined to healthcare environments but can also persist in diverse ecological niches.

The prevalence of *A. baumannii* in the river ecosystem could be attributed to several factors. Effluents from hospitals, pharmaceutical industries, and untreated sewage discharge contribute to the introduction of antibiotic-resistant bacteria into water bodies. Additionally, the ability of *A. baumannii* to survive in harsh conditions, form biofilms, and acquire resistance determinants through horizontal gene transfer enhances its persistence in aquatic environments. River ecosystems with high anthropogenic activities serve as reservoirs and conduits for the dissemination of antimicrobial resistance (AMR), increasing the risk of human exposure and potential outbreaks.

The phylogenetic trees of isolates D-1 and D-2 show close relationships with *Acinetobacter* species and *Acinetobacter baumannii* strain. Short branch lengths and strong bootstrap support indicate a high degree of genetic similarity within the *Acinetobacter* genus based on 16S rRNA gene sequences.

When compared with other studies on AMR in Indian rivers, the findings align with previous reports indicating the widespread presence of multidrug-resistant bacteria in major water bodies. For instance, studies on the Ganges and Yamuna rivers have documented high levels of antibiotic-resistant Enterobacteriaceae and non-fermenting Gram-negative bacteria, underscoring the role of river systems in AMR dissemination. Similar trends have been observed in studies from the Mithi River in Mumbai and the Cauvery River in South India, which reported the occurrence of resistant *Acinetobacter* species, indicating that pollution and urban wastewater significantly contribute to the AMR burden in Indian rivers (Buelow *et al*., 2021).

Continuous monitoring of environmental samples for the presence of *A. baumannii*, combined with comprehensive genetic characterization, is crucial for assessing the potential risks to public health. Environmental surveillance can provide early warnings of emerging strains or resistance patterns that may impact clinical settings. Furthermore, understanding the genetic and ecological factors influencing the survival and spread of *A. baumannii* in the environment can inform strategies to mitigate its impact and prevent potential outbreaks (Buelow *et al*., 2021).

## Conclusion

This study identified antibiotic-resistant bacteria, specifically *Acinetobacter baumannii*, in Daya river water. The detection of strains resistant to ampicillin and ciprofloxacin raises significant concerns about the management and disposal of sewage from the city’s municipal system. Additionally, the study highlights the critical role of advanced bioinformatics tools in elucidating the evolutionary relationships within the *Acinetobacter* genus. These findings highlight the urgent need to monitor and mitigate the spread of AMR in riverine environments. Overall, the findings highlight the Daya River as a reservoir for antibiotic-resistant bacteria, necessitating regular monitoring and effective wastewater management strategies to mitigate the spread of resistance in natural aquatic environments.

Investigating the molecular mechanisms responsible for the observed antibiotic resistance in *A. baumannii* isolates, particularly through whole-genome sequencing and analysis of resistance genes and their regulatory elements, is also vital. Finally, implementing public health measures to reduce water contamination from industrial and human waste will be critical in preventing the spread of antibiotic-resistant bacteria in the environment.

## Data Availability

All data produced in the present study are available upon reasonable request to the authors

## Acknowledgments

The corresponding author is thankful to Raiganj University & Ravenshaw University providing facility of research. We extend our sincere thanks to Biokart India Pvt. Ltd. for their technical assistance.

## Ethical approval

Not applicable.

## Data availability statement

On reasonable request, the corresponding author may disclose any data created and/or analyzed as part of the current work.

